# Detection of SARS-CoV-2 using non-commercial RT-LAMP reagents and raw samples

**DOI:** 10.1101/2020.08.22.20179507

**Authors:** Alisa Alekseenko, Donal Barrett, Yerma Pareja-Sanchez, Rebecca J Howard, Emilia Strandback, Henry Ampah-Korsah, Urška Rovšnik, Silvia Zuniga-Veliz, Alexander Klenov, Jayshna Malloo, Shenglong Ye, Xiyang Liu, Björn Reinius, Simon Elsässer, Tomas Nyman, Gustaf Sandh, Xiushan Yin, Vicent Pelechano

## Abstract

Abstract

RT-LAMP detection of SARS-CoV-2 has been shown as a valuable approach to scale up COVID-19 diagnostics and thus contribute to limiting the spread of the disease. Here we present the optimization of highly cost-effective in-house produced enzymes, and we benchmark their performance against commercial alternatives. We explore the compatibility between multiple DNA polymerases with high strand-displacement activity and thermostable reverse transcriptases required for RT-LAMP. We optimize reaction conditions and demonstrate their applicability using both synthetic RNA and clinical patient samples. Finally, we validated the optimized RT-LAMP assay for the detection of SARS-CoV-2 in raw nasopharyngeal samples from 184 patients. We anticipate that optimized and affordable reagents for RT-LAMP will facilitate the expansion of SARS-CoV-2 testing globally, especially in sites and settings with limited economic resources.

## Introduction

The ongoing SARS-CoV-2 pandemic has a tremendous impact on society, approaching one million casualties worldwide and exposing the vulnerability of our globalised world to the spread of infectious disease. Extensive testing and isolation of confirmed cases has been proposed as a viable strategy to limit the spread of SARSCoV-2 (1–3). Consequently, many methods for SARS-CoV-2 ribonucleic acid detection have been developed (reviewed in (4)). The gold standard for RNA detection is reverse transcription quantitative PCR (RT-qPCR), requiring specialized equipment of limited availability in many contexts *(e.g*. real time thermocyclers). Due to their intrinsic simplicity and high sensitivity, isothermal detection methods are increasingly appreciated as well suited for point-of-care (POC) testing (5), and among them LAMP (loop-mediated isothermal amplification) is an attractive approach (6,7). Early on during the COVID-19 pandemic many researchers, including ourselves, worked to adapt and optimize RT-LAMP detection methods for SARS-CoV-2 diagnostics (8–10). This initial work quickly expanded with optimized oligonucleotide designs and simplified purification systems (11–13). The specificity of RT-LAMP has been improved by applying CRISPR based detection (14,15) or high-throughput sequencing to confirm amplicon sequences and to facilitate mass usage (16,17). Simplified isothermal approaches have the potential to facilitate SARS-CoV-2 diagnosis and thus contribute to the efforts against the COVID-19 pandemic. This is especially important when considering that frequent testing, even at a lower sensitivity, can contribute to a decrease in spread of the disease (1–3).

However, access to reliable and affordable molecular biology reagents remains a problem. Even as the production and distribution has significantly increased since the start of the crisis, the need for testing to control disease spread and allow reopening of society keeps raising the demand. This is especially problematic in countries with limited distribution channels and a lack of economic resources to afford massive testing. To make molecular testing more accessible and allow its widespread implementation, multiple approaches have been developed. For example, it has recently been shown how a simplified RT-qPCR reaction for SARS-CoV-2 detection is feasible using only one enzyme (17,18), or even how it is possible to use crude enzymes from lyophilized bacteria for LAMP (20). Here we aim to contribute to the global effort to generate affordable SARS-CoV-2 tests based on in-house produced enzymes and thus democratize their use. We established protocols for simple production of DNA polymerase with high strand-displacement activity (21) and optimized their reaction conditions for RT-LAMP. We explore their compatibility with thermostable reverse transcriptases and provide a complete inhouse reagent mix for fluorescent or colorimetric detection of SARS-CoV-2 RNA. We then benchmarked the optimized reaction mix with multiple commercial alternatives and RT-LAMP primer sets. Finally, we tested the ability of the produced reagents to correctly detect SARSCoV-2 presence directly in non-purified clinical nasopharyngeal samples from 184 patients.

## Results

### Purification and optimization of alternative LAMP enzymes

As a starting point to optimize alternative reagents for LAMP testing, we used the thermophilic strand-displacing polymerases developed by the Ellington Lab (21). We used both Bst LF from *Geobacillus stearothermophilus* as well as the chimeras v5.9 and v7.16 from Bst LF and Klentaq *(Thermus aquaticus)* which displayed higher thermal stability (21). We performed simple enzyme expression and purification to enable effortless production of good quality reagents for hundreds of thousands of tests (see methods). To validate the performance of the synthesized enzymes in SARS-CoV-2 RT-LAMP detection, we first benchmarked their ability to amplify *in vitro* produced SARS-CoV-2 RNA fragments using the iLACO primer set that we previously developed (8) (Fig 1).

**Figure 1.**
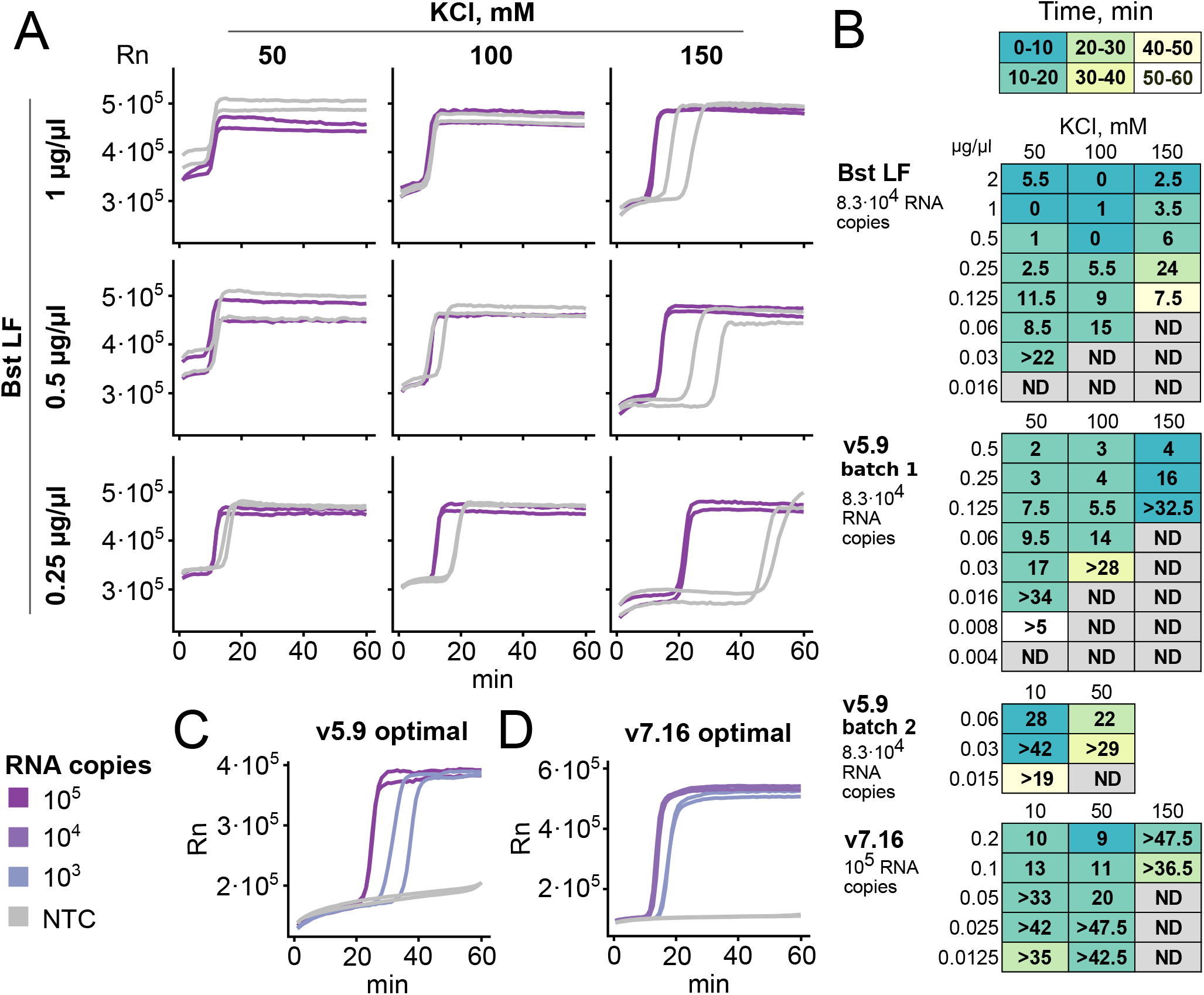
Optimization of enzyme amount and buffer composition for RT-LAMP. All experiments shown were set up with iLACO primers and run for 1 hour at 65 °C in a thermocycler, tracked by either SYBR Green I (Bst LF, v5.9) or Eva Green (v7.16) fluorescence. **A**. Example of RT-LAMP optimization varying Bst LF amount and the KCl concentration. Positive control synthetic RNA (83000 copies) and no template control (each in duplicate) were assayed. **B**. Summary of optimization of enzyme amount and KCl concentration for Bst LF, v5.9, and v.7.16. Numbers indicate the time difference (in minutes) between the latest-amplifying positive control replicate and the earliest-amplifying negative control replicate (i.e. high numbers represent clear difference between positive and negative controls). Color indicates the required time to detect the latest-amplifying positive control (i.e. time to positive detection). “>…” indicates that the negative control did not amplify within 1 hour. ND indicates no amplification of either positive or negative control. Two batches of v5.9 with slightly different activity were assayed, and batch 2 was used for further experiments. **C-D**. Optimal conditions determined for v5.9 (0.03 µg/µl in ThermoPol buffer) and v7.16 (0.025 µg/µl in isothermal amplification buffer). 83000 and 830 copies of RNA were used with v5.9, 10000 and 1000 with v7.16.

We tested the intrinsic reverse transcriptase (RT) activity of all enzymes, and compared their ability to efficiently amplify and detect the synthetic RNA fragment with or without supplementing the reaction with a commercial thermostable RT enzyme (Superscript IV, SSIV). Addition of a thermostable RT was essential to enhance the performance of all tested enzymes (not shown), thus, we carried out further optimization in the presence of SSIV. To facilitate quantitative comparison between samples, we performed all the optimizations using continuous fluorescence detection. However, to increase throughput and avoid to use expensive equipment, end-point measurement of fluorescence or colorimetric detection can also be applied (8–13,17,22).

We optimized the composition of the reaction buffer and the amount of strand displacing DNA polymerase used (Fig. 1A-B). For a fixed template amount we minimized the time to detection (color in Fig. 1B) and maximized the time between true positive and false positive (spurious amplification, number in Fig. 1B). Non-specific spurious LAMP amplification is a known phenomenon that can lead to false positive detection and is templated by the primers themselves (23). In some cases the amplification of spurious non-specific sequences can be detected by performing a melting curve analysis (24). However, to avoid false positives, reaction conditions and components should be optimized to maximize sensitivity while minimizing spurious amplification. Variation of any component of the reaction *(e.g*., oligos, enzyme, buffer composition…) as well as the physical reaction conditions (e.g. temperature, time…) can affect the spurious amplification. For example, while increased enzyme concentration decreases time to detection, it also increases false positive amplification (Fig. 1B-C). We observed that the optimal concentration of KCl varied between the enzymes tested. 150 mM KCl was optimal for Bst LF, 50 mM for v7.16, and 10 mM for v5.9. Based on previous literature, we tested the capacity of betaine (25), guanidine (26) and DMSO (27) to increase sensitivity and reduce nonspecific amplification. Unfortunately, in our hands none of these strategies improved sensitivity. It is important to note that even the dye used for fluorescent detection can alter the measured RTLAMP sensitivity and specificity and thus should also be accounted for optimization. In our conditions, Eva Green dye provided a higher dynamic range than SYBR Green I, when measuring difference in fluorescence before and after RT-LAMP reaction. We compared the three polymerases based on the speed of amplification, the specificity, and the range of conditions where the former two parameters are acceptable. We found that v7.16 outperformed v5.9, and both outperformed Bst LF. Amplification curves for LAMP at optimal conditions with v5.9 and v7.16 are shown in figures 1C and 1D. We proceeded with both v5.9 and v7.16 in the following experiments.

### Optimization of alternative RT enzymes compatible with LAMP

Once we optimized the conditions for efficient strand-displacement DNA activity required for LAMP, we decided to explore alternatives to the commercial thermostable RT enzymes. First, we explored the use of a thermostable synthetic reverse transcriptase, RTX, developed by the Ellington lab (28), which can also be used for direct RT- qPCR of SARS – CoV-2 (18). Supplementing either v5.9 or v7.16 with different concentrations of RTX did however not improve their ability to detect SARS-CoV-2 RNA (Fig. 2A). Addition of high concentrations of RTX lead to abnormal amplification curves, while lower concentration did not improve the RT-LAMP activity of v5.9 and v7.16. Similar results were obtained using both the RTX versions containing or excluding the proofreading domain (Fig. S1AD). Importantly, the absence of improvement in the LAMP reaction was not due to lack of RT activity of these enzymes, as demonstrated by standard RT-qPCR (Fig. S1E). Thus, we concluded that RTX is not compatible with RTLAMP.

In addition to RTX, we aimed to explore other noncommercial alternatives. MashUp-RT is based on FeLV-RT, which is intrinsically more accurate than MMLV-RT (29) and contains mutations which further increase thermostability and fidelity (https://pipettejockey.com/). We expressed MashUp-RT and tested its ability to enhance RT-LAMP reaction in combination with v5.9 and v7.16. The addition of 0.025 µg/µl reaction was enough for a performance identical to, or even better than, that of SSIV (Fig. 2B-C). This shows that MashUp-RT is a thermostable RT enzyme compatible with LAMP that is capable of enhancing RT-LAMP detection sensitivity of SARS-CoV-2 RNA.

**Figure 2.**
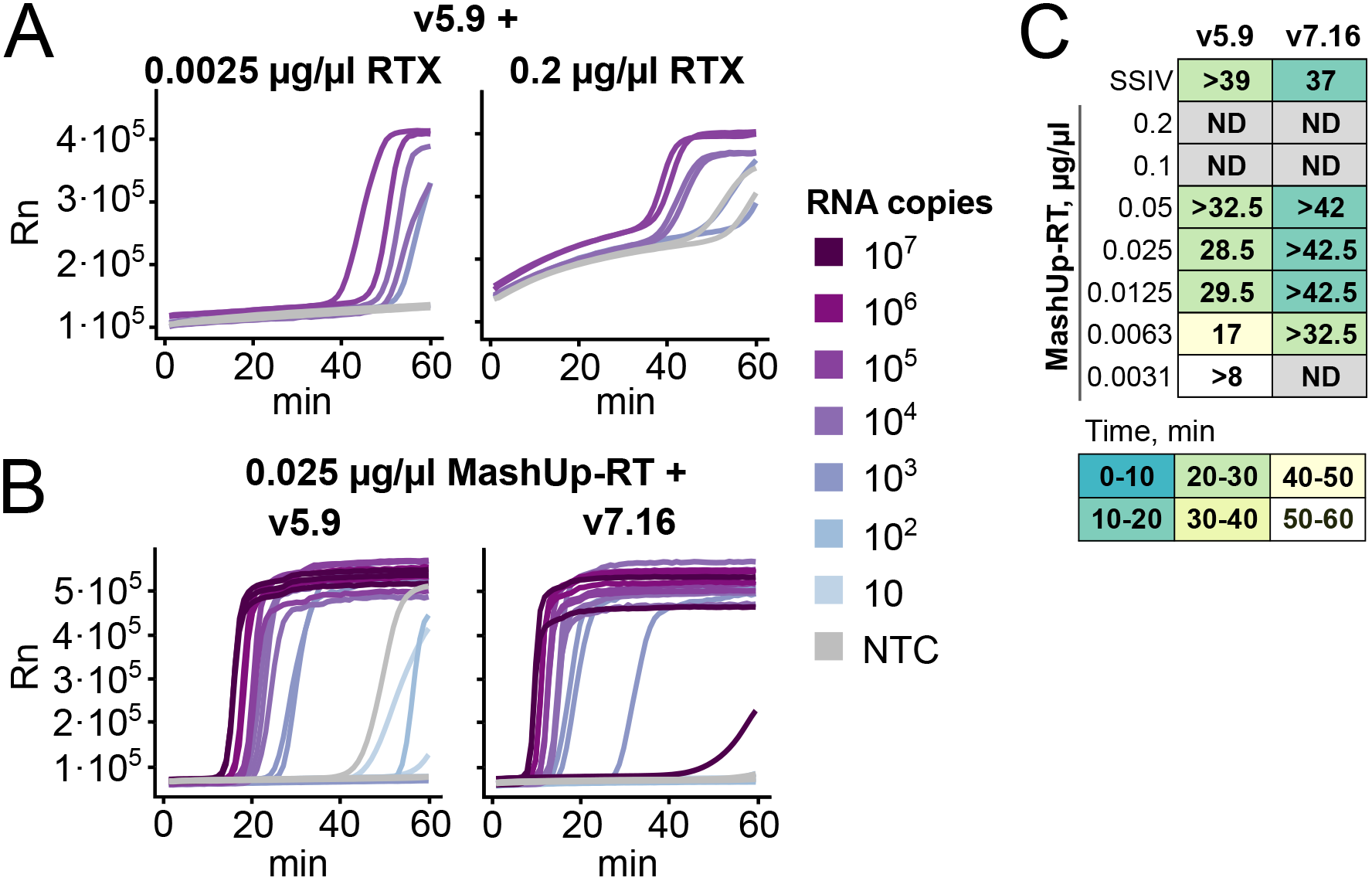
Compatibility of v5.9 and v7.16 polymerases with non-commercial thermostable RT enzymes. All experiments shown were set up with iLACO primers and run for 1 hour at 65 °C in a thermocycler, tracked by Eva Green fluorescence. **A**. Example amplification plots showing the performance of v5.9 with low or high amounts of RTX (duplicates of 83000, 8300, and 830 copies of synthetic RNA as well as non template control (NTC)). **B**. Optimal detection of synthetic SARS-CoV-2 RNA using v5.9 (0.03 µg/µl in ThermoPol buffer) or v7.16 (0.025 µg/µl in IA buffer) supplemented with MashUp-RT as the thermostable reverse transcriptase. Reactions were performed in triplicate. **C**. Optimization of MashUp-RT enzyme amount in combination with v5.9 and v7.16 in their respective optimal conditions (see above for B). 10000 copies of synthetic RNA and NTC were assayed. Time between positive and negative amplification is indicated in minutes and the color indicates time for detection of the positive controls (as in Fig. 1).

### Benchmarking of alternative RT-LAMP enzyme mix and oligo combinations

Once we optimized non-commercial enzyme RTLAMP conditions, we benchmarked their ability to detect SARS-CoV-2 against commercial alternatives. After testing multiple oligo primer sets optimized for SARS-CoV-2 (8,10,12), we decided to focus on the use of iLACO (8) and As1e (12) targeting ORF1ab that worked better in our hands. We compared our optimized enzyme mix with commercial alternatives from multiple providers *(i.e*., WarmStart Colorimetric master mix (NEB), Bst3.0 (NEB) and Saphir Bst2.0 Turbo (Jena Biosciences)) using synthetic RNA (Fig. 3). Although all enzymes were able to detect SARSCoV-2 RNA, we identified clear differences both in sensitivity and background level. In particular, the performance of the RT-LAMP reaction using v7.16 enzyme and As1e primer was comparable to the best commercial alternatives. With a detection speed similar to Bst3.0 and lower spurious amplification. Unexpectedly, in our hands Bst3.0 needed to be supplemented with RT activity to detect SARS-CoV-2 RNA (Fig. S2). The performance of primer sets was also affected by the different enzymes used. For example, v5.9 and Saphir Bst2.0 perform better in combination with iLACO primers, while other enzymes perform better with As1e or similarly with both. Thus we concluded that in-house produced v5.9 or v7.16 in combination with MashUp-RT and iLACO or As1e primers were competitive alternatives for RTLAMP detection of purified RNA.

**Figure 3.**
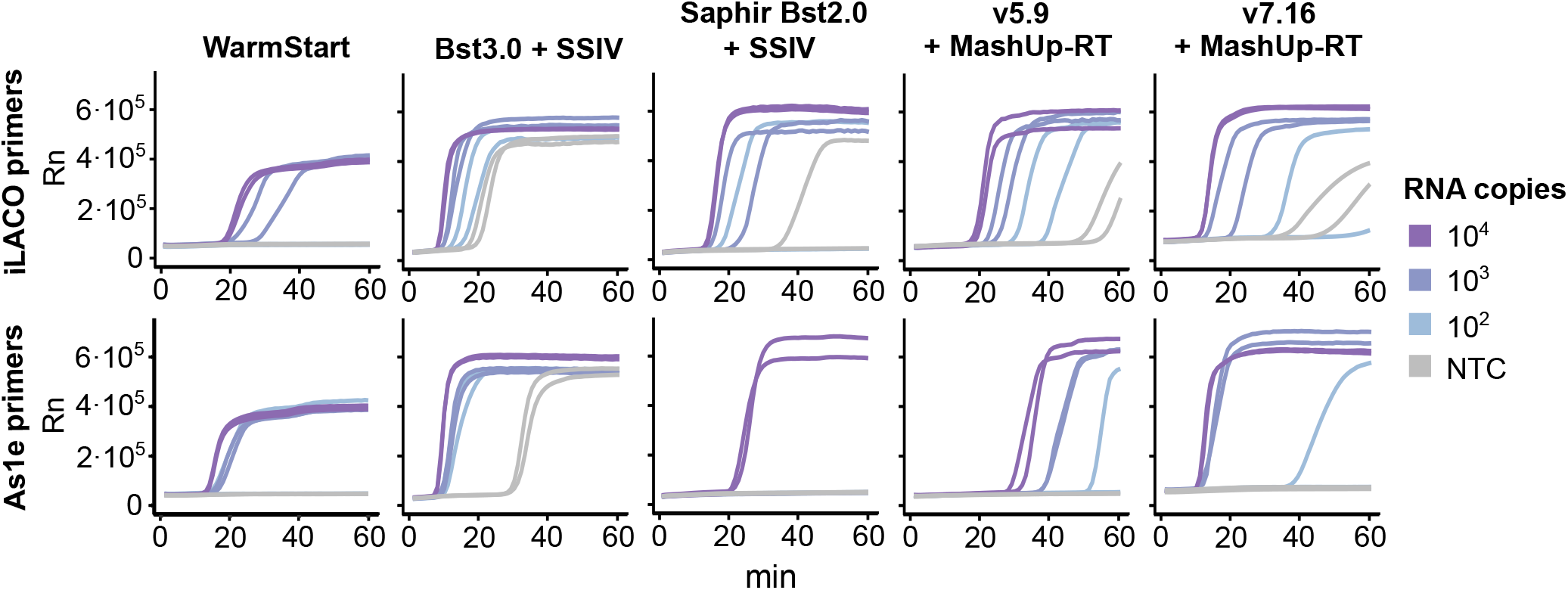
Benchmarking of in-house produced enzymes against commercial alternatives. Synthetic RNA templates (iLACO and As1e) amplified with the corresponding primers and either WarmStart, Bst3.0 with SSIV, Saphir Bst2.0 Turbo with SSIV, v5.9 with MashUp-RT, or v7.16 with MashUp-RT (same conditions for v5.9 and v7.16 as in Fig 2B). All experiments were run for 1 hour at 65 °C in a thermocycler, tracked by Eva Green fluorescence.

### Application of optimized RT-LAMP to clinical nasopharyngeal samples

As we and others have already shown, the utility of RT-LAMP for the detection of SARS-CoV-2 from patient purified RNA (8–11,17), we wanted to explore now the ability of our in-house enzyme mix to detect SARS-CoV-2 also in non-purified samples (11,17).

We first explored its compatibility with common virus transport media commonly used for collection of nasopharyngeal swabs: Virocult (MWE), Sigma Transwab (MWE), eSwab (Copan) and Beaver (BEAVER biomedical) (Fig. 4A). Three of the four tested media (Virocult, Transwab and eSwab) were compatible with both v7.16 and v5.9 reactions when adding 10% of the reaction volume. However, the Beaver media required additional dilution to avoid inhibition and was compatible only with v5.9. That limits the total amount of patient sample that could be used per reaction and thus decreases the overall sensitivity.

**Figure 4.**
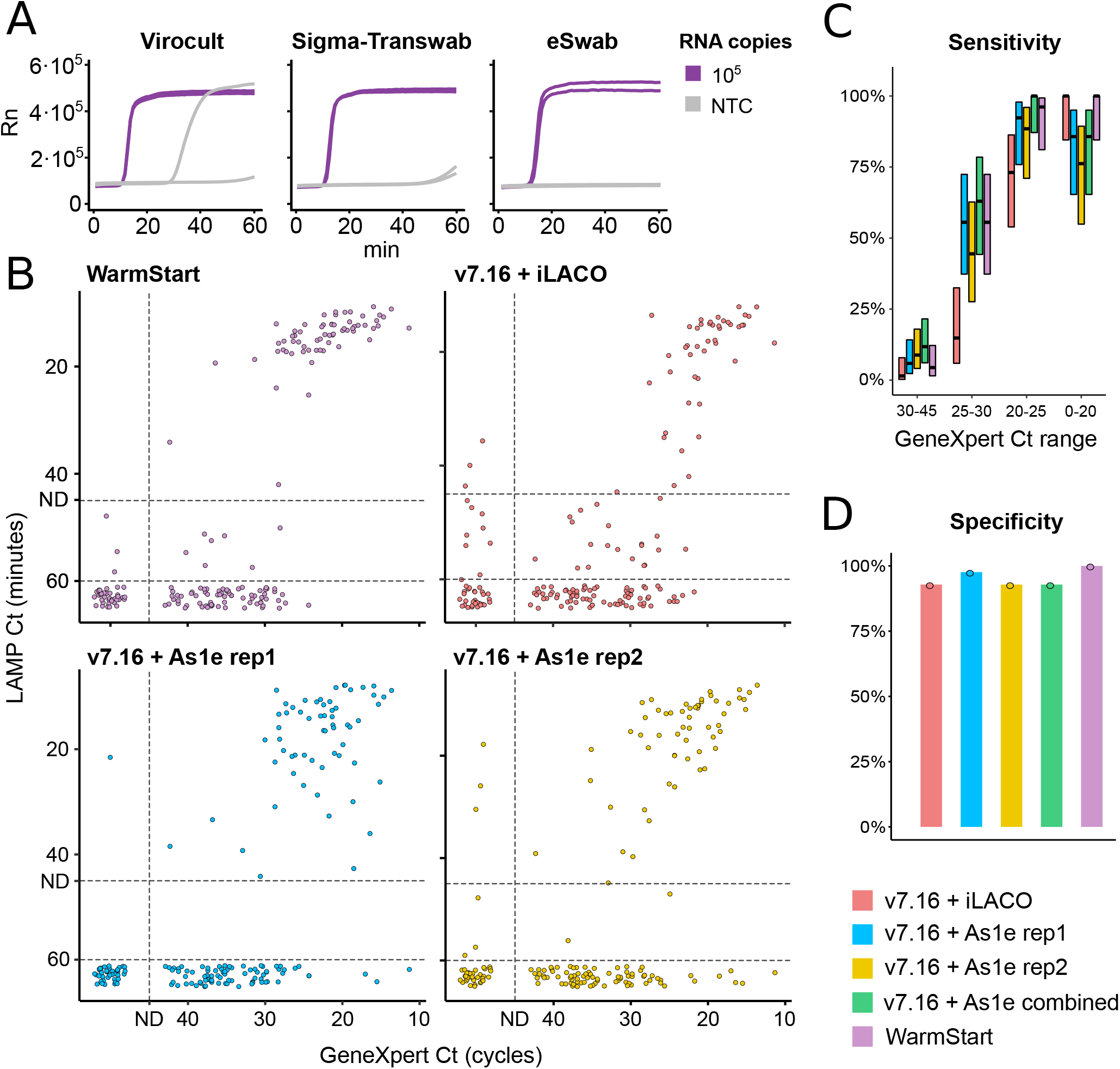
Applicability of RT-LAMP on raw nasopharyngeal samples. A. Effect of common virus transport media on RT-LAMP amplification with v7.16 and MashUp-RT (same conditions and same experiment as in Fig. 2B). Different transport media were added at 10% of the reaction volume. **B**. Comparisons of RT-LAMP Ct (minutes) and GeneXpert RT-qPCR Ct (cycles) for 184 clinical samples. The developed v7.16+RT-MashUP reaction mix was tested with iLACO (red) or As1e primer sets (blue/yellow). As1e primer set was also tested with commercial WarmStart Colorimetric master mix (violet). ND designates the thresholds for calling positives (see methods). **C**. Reaction sensitivity according to SARS-CoV-2 abundance as determined by GeneXpert. When technical replicates of As1e were performed and considered together, sensitivity improved (in green, positives were called when at least one replicate was identified as positive). Number of samples in each category is indicated, and 95% confidence intervals are shown (Wilson’s binomial CI) (17). **C**. Reaction specificity, as % of samples considered negatives by RT-qPCR that were also negative by RT-LAMP.

Having established the reaction conditions, we used 184 nasopharyngeal samples collected in Virocult, TransSwab and eSwab transport media that have been previously analyzed using GeneXpert SARS-CoV-2 detection (Cepheid) at the Karolinska University Hospital (Sweden). We selected 142 positive samples across all GeneXpert Ct ranges and 42 samples determined to be negative for SARS-CoV-2. We used the developed v7.16 + MashUp-RT reaction mix in combination with either iLACO or As1e primer sets (Fig. 4B-D and Supplementary table 1). While both primer sets were successful in detecting samples with high viral load, the As1e primers showed better overall sensitivity. Performing technical duplicates demonstrated that the sensitivity was further increased if a sample was called as positive when at least one replicate had amplified (Fig. 4C). Unfortunately, in all cases patient samples with low-to-medium content of SARS-CoV-2 RNA (higher GeneXpert Cts) were often misclassified as false negatives. Finally, we compare the results obtained with our in-house reagents with state-of-the-art commercial reagents (Warmstart Colorimetric master mix) and demonstrate that they have comparable performance in terms of specificity and sensitivity. In summary, we have shown how in-house optimized enzyme mixes offer a high quality economic alternative to current commercial products.

## Discussion

Here we have optimized and validated the production and application of alternative noncommercial reagents for simplified RT-LAMP based SARS-CoV-2 detection. We demonstrate ability to correctly identify COVID-19 positive patients using raw nasopharyngeal swabs in common virus transport media. And finally, we show that even omitting RNA purification, RTLAMP can easily detect samples with high to medium SARS-CoV-2 RNA content. This is especially important when considering that frequent testing, even at a lower sensitivity, can help to effectively decrease disease spread (1–3). During this optimization process we obtained some general recommendations that can facilitate the application of RT-LAMP in different settings. In addition to the initial benchmarking with alternative RT-qPCR approaches, it is important to routinely control for spurious amplifications or cross contaminations. For example, it is common to observe variation in the quality and specificity of different oligonucleotide batches (even from the same provider). In some cases oligonucleotide batches can contain a trace amount of target regions from the SARS-CoV-2 genome due to oligo cross contamination during oligonucleotide synthesis or handling. Thus reactions with negative water controls should always be performed in parallel to ensure the specificity of the test. Additionally, the activity of each new batch of enzymes and reagents should first be calibrated with a known serial dilution of target RNA. Although direct RT-LAMP in crude samples can detect most cases with medium to high SARS-CoV-2 viral load, to bring it to RT-qPCR detection sensitivity it is still necessary to perform at least some level simple sample purification and RNA concentration (9–11,15). In addition, even if the use of virus transport media can be tolerated by RT-LAMP, less complex media, or no media at all, might be more advantageous (30). Our work also highlights the importance of controlling for the presence of salt and other components that may affect RT-LAMP sensitivity and specificity. In summary, we hope that this work will facilitate the implementation of affordable RT-LAMP SARSCoV-2 diagnostic in settings where the use of commercial reagents is not possible. This could decrease the total cost per test in order to facilitate routine testing of large portions of the population.

## Data Availability

Data provided as supplementary table.

## Acknowledgements

Acknowledgments

We thank Andy Ellington and Alexander Klenov (https://pipettejockey.com/) for sharing regents. We thank Björn Högberg, Sten Linnarsson, Michael Knop, Simon Anders, Uwe Sauer and the whole SciLifeLab community for discussion and support.

## Ethics statement

We used pseudo-anonymized surplus material previously collected for clinical diagnostics of SARSCoV-2. This is in accordance with the Swedish Act concerning the Ethical Review of Research Involving Humans, which allows development and improvement of diagnostic assays using patient samples which were collected to perform the testing in question. Additional ethical approval for RT-LAMP diagnosis was obtained by the appropriate Swedish Authority (Dnr 2020–01945, Etikprövningsnämnden).

## Methods

### Expression and purification of strand displacing polymerases

Displacement polymerases assayed for RT-LAMP polymerase activity included the *Geobacillus stearothermophilus* exonuclease-deficient family-A polymerase large fragment (Bst-LF), and its ‘v5.9’ and ‘v7.16’ derivatives previously reported by Ellington and colleagues to incorporate functional properties of the related Klentaq polymerase from *Thermus aquaticus* (21). Prior to functional characterization, hexahistidine (H6)-tagged variants were screened for expression efficiency in various vectors, including pTetA (constructs kindly provided by Andrew D Ellington), pET-16b and pET-28a (synthesized for this work). Constructs in pET-16b contained an ASRGS-H6 followed by the polymerase sequence, inserted between NcoI and BamHI sites in the multiple cloning region. Similarly, constructs in pET-28a were engineered by inserting the polymerase between NdeI and XhoI, 3’ to the incorporated GSS-H6-SSG and thrombin cleavage sequences.

Polymerase expression and purification protocols were adapted from that of Milligan and colleagues (21). Briefly, cells were cultured overnight at 37° C, followed by inoculation (1:200) into fresh 2xYT media with suitable antibiotic (100 mg/L ampicillin for constructs in pTetA and pET-16b, 50 mg/L kanamycin for pET-28a). After cells reached an optical density (OD600) of 0.6– 0.8, expression was induced (200 µg/L tetracycline-HCl in 70% ethanol for pTetA, 100 µM isopropyl-β-D-1-thiogalactopyranoside for pET-16b or pET-28a) and cells grown for an additional 3–7 hours at 30–37° C before harvesting. Pellets were resuspended, sonicated, and cleared by ultracentrifugation (30 min at 40,000 × g, 4° C) in buffer A (20 mM Tris pH 7.4, 300 mM NaCl, 0.1% Tween-20, 10 mM imidazole) supplemented with 500 mg/L lysozyme and EDTA-free protease inhibitors. As an initial purification step, lysate supernatants were then heated to 65° C for 20 min with gentle shaking (400 rpm), then again cleared by ultracentrifugation (20 min at 20,000 × g, 4° C). Clarified supernatants were purified by Ni^2+^ affinity (HisPur Ni-NTA resin), eluting in buffer A with 250 mM imidazole. Positive elution fractions were pooled and concentrated in storage buffer (10 mM Tris pH 7.4, 100 mM KCl, 1 mM DTT, 0.1 mM EDTA, 0.5% Tween-20, 0.5% Triton-X100, 50% glycerol), then aliquoted, flash-frozen in liquid nitrogen, and stored at –80° C. All constructs yielded at least 3 mg purified protein per liter liquid culture, sufficient to amplify thousands of individual RT-LAMP samples at negligible consumables cost.

### Expression and Purification of thermostable reverse transcriptases

The MashUp-RT plasmid (kindly provided by https://pipettejockey.com/) was transformed into BL21 (DE3) T1R pRARE2 expression strain and plated on L-Broth (LB) agar (Formedium, Norfolk, UK) plate containing 50 µg/mL kanamycin. Overnight cultures were inoculated with fresh transformants and grown at 37 °C at 175 RPM in LB medium (Formedium, Norfolk, UK) containing kanamycin (50 µg/mL). Subsequently, the overnight culture was diluted into 1.5 L fresh LB medium supplemented with kanamycin and catabolite repression buffer (25% glycerol, 25% glucose, 1 mM MgSO4 and 0.1 mM MnCl2) and the cultures were grown at 37 °C in the LEX system (LEX-48, Epiphyte Three Inc., Toronto, Canada). At OD600 0.9, the cultures were down-tempered to 18 °C before protein expression was induced with 0.5 mM IPTG and the culture was grown additionally for 24 h at 18 °C. The cells were harvested by centrifugation at 4500 × *g* for 10 min at 4 °C and resuspended in MashUp-RT IMAC lysis buffer (25 mM Tris-HCl, 300 mM NaCl, 10% glycerol, 40 mM imidazole, 0.5% Triton X-100, pH 8) supplemented with 1 mg/mL lysozyme and one tablet Complete EDTA-free protease inhibitor cocktail (Roche Diagnostics GmbH, Mannheim, Germany) per 1.5 L culture. The resuspended cell pellets were stored at –80 °C.

The pET2 vector containing the coding sequence of RTX (reverse transcriptase) and RTX^exo-^(RTX lacking exonuclease activity), kindly provided by Andy Ellington, were transformed into BL21(DE3) T1R pRARE2 expression strain and plated on LB plates containing carbenicillin (50 µ g/mL) a n d chloramphenicol (34 µg/mL). Terrific Broth (TB) medium (Merck, Darmstadt, Germany) supplemented with carbenicillin (100 µg/mL) and chloramphenicol (34 µg/mL), 8 g/L glycerol and 0.4% glucose were inoculated with fresh transformants and grown overnight at 30 °C at 175 RPM. The overnight cultures were diluted into 3 L (RTX) or 1.5 L (RTX^exo-^) fresh TB medium supplemented with 8 g/L glycerol and carbenicillin (50 µg/mL). The cultures were grown at 37 °C in the LEX system and the OD600 was measured periodically until OD600 2 at which the temperature was set to 18 °C. Protein expression was induced at approximately OD600 3 with 1 mM (RTX) or 0.5 mM (RTX^exo-^) IPTG and the cultures were grown additionally for 24 h. The cells were sedimented by centrifugation at 4500 × *g* for 10 min at 4 °C. The cell pellets were then resuspended in RTX IMAC lysis buffer (100 mM HEPES, 500 mM NaCl, 10% glycerol, 10 mM imidazole, 0.5 mM TCEP, pH 8.0) and stored at –80 °C.

The resuspended cells from each culture were thawed at room temperature and subsequently lysed by pulsed sonication (4s/4s 4 min, 80% amplitude, Sonics Vibracell-VCX750, Sonics & Materials Inc., Newtown, CT, US). The crude cell lysates were centrifuged for 20 min at 49000 ×*g* at 4 °C. For RTX and RTX^exo-^ the supernatants were incubated for 20 min at 65 °C and subsequently clarified by centrifugation at 49000 × *g* for 20 min at 4 °C. The supernatants were filtered through a 0.45 µm filter (Corning bottle-top vacuum filter, 0.45 µm, Corning, NY, USA), and then loaded onto a preequilibrated 1 mL (MashUp-RT) or 5 mL (RTX and RTX^exo-^) HisTrap HP column (Cytiva, Little Chalfont, UK). All proteins were purified using ÄKTAXpress systems (Cytiva, Little Chalfont, UK). For MashUp-RT, the IMAC column was washed with 20 column volumes (CV) of MashUp-RT IMAC lysis buffer (25 mM Tris-HCl, 300 mM NaCl, 10% glycerol, 40 mM imidazole, 0.5% Triton X-100, pH 8) and the bound protein was eluted with 2 CV of 0–6% MashUp-RT IMAC elution buffer (25 mM Tris-HCl, 300 mM NaCl, 10% glycerol, 500 mM imidazole, 0.5% Triton X-100, pH 8), 2 CV of 6–10% elution buffer and finally 24 CV of 10–100% elution buffer. For RTX and RTX^exo-^, the IMAC column was washed with 20 CV of wash buffer 1 (20 mM HEPES, 500 mM NaCl, 10% glycerol, 10 mM imidazole, 0.5 mM TCEP, pH 7.5) and 20 CV of wash buffer 2 (20 mM HEPES, 500 mM NaCl, 10% glycerol, 50 mM imidazole, 0.5 mM TCEP, pH 7.5) and thereafter eluted with 5 CV of RTX IMAC elution buffer (20 mM HEPES, 500 mM NaCl, 10% glycerol, 500 mM imidazole, 0.5 mM TCEP, pH 7.5). For RTX and RTX^exo-^ the proteins were further purified on a HiLoad 16/60 Superdex 200 preparative grade column (Cytiva, Little Chalfont, UK) equilibrated with gel filtration buffer (20 mM HEPES, 300 mM NaCl, 10% glycerol, 0.5 mM TCEP, pH 7.5). Elution fractions were analyzed by SDS-PAGE before pooling and the fractions containing the target proteins were concentrated with Amicon Ultra-15 concentration filter units (50 kDa cut off, Millipore, Burlington, MA, USA) at 5000 × *g*, 4 °C. For MashUp-RT, the elution buffer was exchanged to buffer A (50 mM Tris, 150 mM NaCl, 1 mM DTT, 1 mM EDTA, 0.01% Triton X-100, pH 7.5) on a PD-10 column (Cytiva, Little Chalfont, UK) and the purified MashUp-RT was then diluted in buffer B (50 mM Tris, 150 mM NaCl, 1 mM DTT, 1 mM EDTA, 0.01% Triton X-100, 60% glycerol, pH 7.5) to obtain the MashUp-RT storage buffer (50 mM Tris, 150 mM NaCl, 1 mM DTT, 1 mM EDTA, 0.01% Triton X-100, 50% glycerol, pH 7.5). For RTX and RTX^exo-^, the elution buffer was exchanged to buffer C (50 mM Tris-HCl, 50 mM KCl, 0.1% Tween-20, 10% glycerol, pH 8.0) on a PD-10 column (Cytiva, Little Chalfont, UK) and the purified protein was then diluted in buffer D (50 mM Tris-HCl, 50 mM KCl, 0.1% Tween-20, 60% glycerol, pH 8.0) to obtain the RTX storage buffer (50 mM Tris-HCl, 50 mM KCl, 0.1% Tween-20, 50% glycerol, pH 8.0). The protein samples were flash-frozen in liquid nitrogen and stored at –80 °C. Working aliquots were stored at –20 °C.

### RT-LAMP primer design and positive control

We used either the iLACO (8) or As1e (12) primers as previously described.

The iLACO primers are: F3

(CCACTAGAGGAGCTACTGTA), B3

(TGACAAGCTACAACACGT), FIP (AGGTGAGGGTTTTCTACATCACTATATTGGAACAAGCAAATTCTATGG), BIP

(ATGGGTTGGGATTATCCTAAATGTGTGCGAGCAAGAACAAGTG), LF

(CAGTTTTTAACATGTTGTGCCAACC) and LB

(TAGAGCCATGCCTAACATGCT). The iLACO primers are: As1_F3 (CGGTGGACAAATTGTCAC), As1_B3 (CTTCTCTGGATTTAACACACTT), As1_LF

(TTACAAGCTTAAAGAATGTCTGAACACT), As1_LB

(TTGAATTTAGGTGAAACATTTGTCACG), As1_FIP

(TCAGCACACAAAGCCAAAAATTTATCTGTGCAAAGGAAATTAAGGAG), As1_BIP

(TATTGGTGGAGCTAAACTTAAAGCCCTGTACAATCCCTTTGAGTG), As1e_FIP

(TCAGCACACAAAGCCAAAAATTTATTTTTCTGTGCAAAGGAAATTAAGGAG) and As1e_BIP

(TATTGGTGGAGCTAAACTTAAAGCCTTTTCTGTACAATCCCTTTGAGTG).

All oligonucleotides were purchased from Thermo Fisher Scientific with standard desalting and dissolved in nuclease free water upon arrival.

We generated synthetic fragments of SARS-CoV-2 RNA (200–300 bp) by *in vitro* transcription of PCR fragments containing part of SARS-CoV-2 Sequence and a T7 promoter. For iLACO we amplified the PCR product from the viral genome using T7_FW_iLACO

(TAATACGACTCACTATAGGGTCAATAGCCGCCACTAGA) and RV_iLACO (AGAAACGGTGTGACAAGCTAC). For As1/As1e we used T7-HMS1_FW

(TAATACGACTCACTATAGGGTGCTTGTGAAATTGTC GGTGGA) and HMS1_rv

(GCTTTTAGAGGCATGAGTAGGC). RNA was produced using the TranscriptAid T7 High Yield Transcription Kit (Thermo Fisher Scientific), DNase-treated using the TURBO DNA-*free* Kit (Invitrogen), purified using Ampure XP beads (Beckman Coulter) and quantified using Qubit (Thermo Fisher Scientific).

### RT-LAMP reaction conditions

RT-LAMP assays were assembled in a total reaction volume of 10 (for optimizations and benchmarking) or 20 (for patient sample tests) µl in MicroAmp Fast Optical 96 well reaction plates (Thermo Fisher Scientific) on ice. For in-house produced enzymes, each 10 µl reaction consisted of: 1 µl of 10X LAMP primer mix (2 µM F3, 2 µM B3, 16 µM FIP, 16 µM BIP, 4 µM LF and 4 µM LB), 1 µl of 10X buffer (one of ThermoPol Reaction Buffer, Isothermal Amplification Buffer or II), 0.5 µl 20X fluorescent dye (either Eva Green (Biotium) or SYBR Green I (Invitrogen), 0.6 µl 100 mM MgSO4 (NEB), 1.4 µl of dNTP mix (10mM each, NEB), 0.5 µl of strand-displacing polymerase, 0.5 µl of in-house reverse transcriptase or 0.2 µl SSIV (Thermo Fisher Scientific), 1 µl sample, and nuclease free water (Invitrogen) up to 10 µl. The in-house enzymes were pre-diluted to desired concentration in a buffer containing 10 mM Tris-HCl (pH 7.5) and 25 mM KCl. For 20 µl reactions, all the volumes above were doubled.

10X ThermoPol Reaction Buffer contains 20 mM Tris-HCl, 10 mM (NH4)2SO4, 10 mM KCl, 2 mM MgSO4, 0.1% Triton X-100, pH 8.8. 10X Isothermal Amplification Buffer contains 20 mM Tris-HCl, 10 mM (NH4)2SO4, 50 mM KCl, 2 mM MgSO4, 0.1% Tween 20, pH 8.8, while 10X Isothermal Amplification Buffer II has the same components but more KCl (150 mM). Some experiments were supplemented with additional KCl to test the full range of concentrations.

Reactions containing Bst 3.0 DNA Polymerase (NEB) and Saphir Bst2.0 DNA polymerase (Jena Bioscience) were largely performed as described for the in-house reagents. Isothermal amplification buffer II was used with Bst3.0 and Saphir Bst2.0 Turbo Buffer (Jena Bioscience) was used with Saphir Bst2.0, as recommended by manufacturers. Enzyme amounts were those recommended by manufacturers. When using WarmStart Colorimetric RT-LAMP 2X Master Mix (NEB), 5 µl mix was used together with 1 µl 10X LAMP primer mix, 0.5 µl Eva Green dye, 1 µl of sample, and nuclease free water to a final volume of 10 µl (double the volumes for 20 µl reactions). All reactions were run in a Step One Plus Real time PCR system (Applied Biosystems).

To test the RT activity of RTX and RTX^exo-^, we performed RT-qPCR according to the following protocol. 10 µl RT reactions were set up for each condition containing: either ThermoPol buffer, Isothermal Amplification buffer, or Isothermal Amplification buffer II; 1.4 mM each dNTP; 6 mM MgSO4 (where appropriate); 0.2 µM iLACO-qPCR-rv primer (AGCATGTTAGGCATGGCTCT); specified amount of either RTX or RTX^exo-^, or 4 U/µl SSIV, or no enzyme; 83 million copies of iLACO synthetic RNA template; and nuclease-free water up to 10 µl. RT was performed at 65 °C for 15 min. 1 µl of each RT reaction was used as template in a 10 µl qPCR reaction using 5 µl Fast SYBR Green Master Mix (Applied Biosystems) and 0.5 µ MeachofiLACO_qPCR_fw (TATGGTGGTTGGCACAACAT) and iLACO_qPCR_rv primers. qPCR was performed according to the standard protocol.

### Clinical samples

We obtained 184 nasopharyngeal samples from Karolinska University Hospital, Huddinge (Stockholm) collected between May 20^th^ and June 1^st^ 2020. We picked samples collected in either Sigma-Transwab, Sigma-Virocult or COPAN-eSwab kits. All samples were first analyzed on the GeneXpert Xpress SARSCoV-2 detection system (Cepheid) at the hospital and called as either positive (142 samples) or negative (42 samples) (Table S1). Samples which had at least a determined GeneXpert Ct value for N2 were called as positive, while samples where both GeneXpert Ct values were not determined were called as negative. For RT-LAMP, 50 µl of each sample was heat-inactivated at 95 °C for 15 minutes and stored at –80 °C prior to experiments. Two µl of raw sample were used in 20 µl total reaction volume.

## Supplementary Material

**Figure S1.**
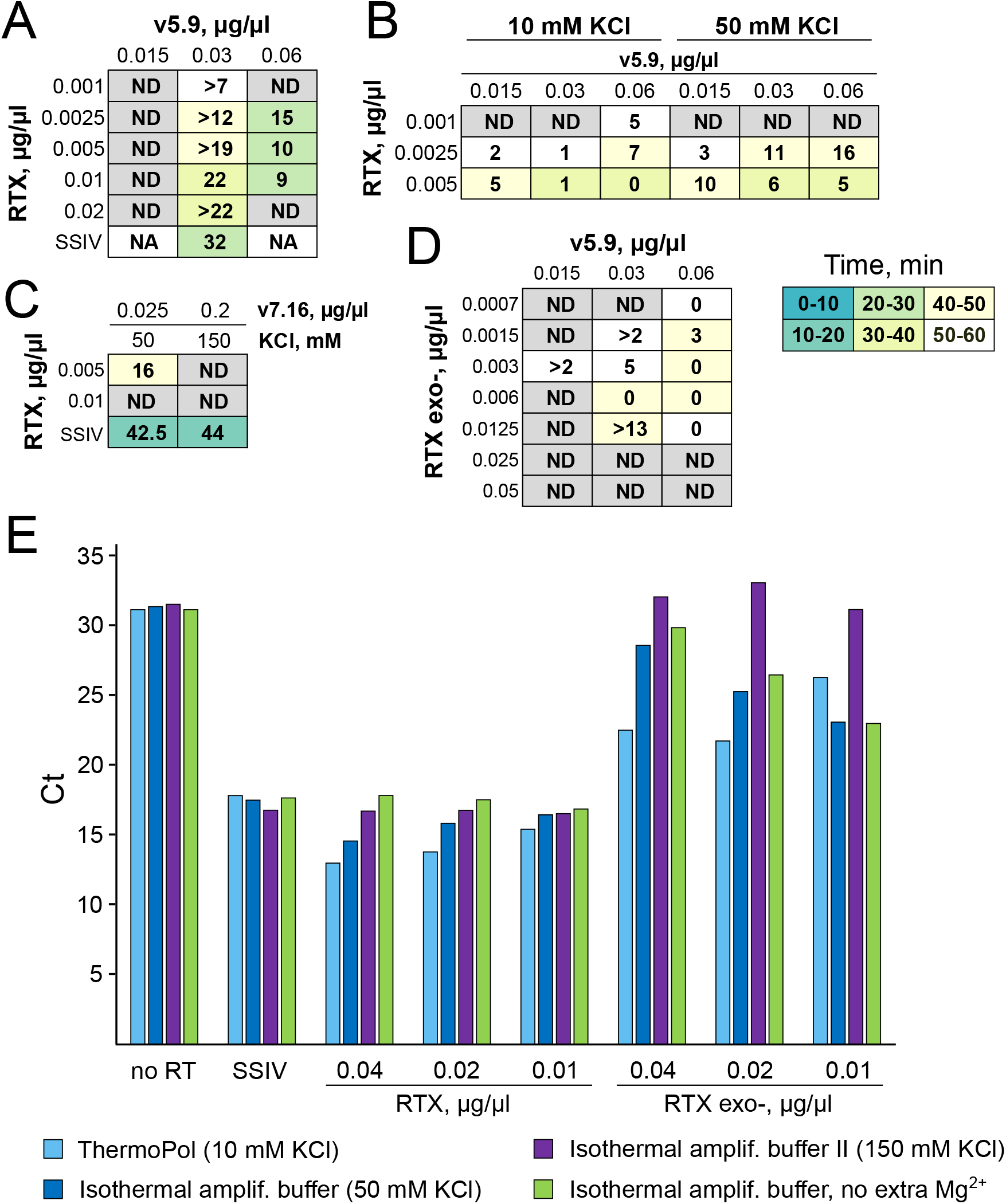
Analysis of RTX performance. Tables showing the performance of RTX together with either v5.9 or v7.16 at different concentrations of the enzymes and with different salt concentrations. The numbers and colors were determined in the same way as in Figures 1 and 2. **A**. 83000 copies of synthetic RNA. **B**. 8300 copies. **C**. 1000 copies. **D**. Table showing performance of RTX^exo-^ together with v5.9 at different enzyme concentrations (8300 copies of synthetic RNA). **E**. Results of RT-qPCR demonstrate that RTX and RTX^exo-^ do possess RT activity, as compared to SSIV (see methods). However, RTX^exo-^has poor performance at higher KCl concentrations.

**Figure S2.**
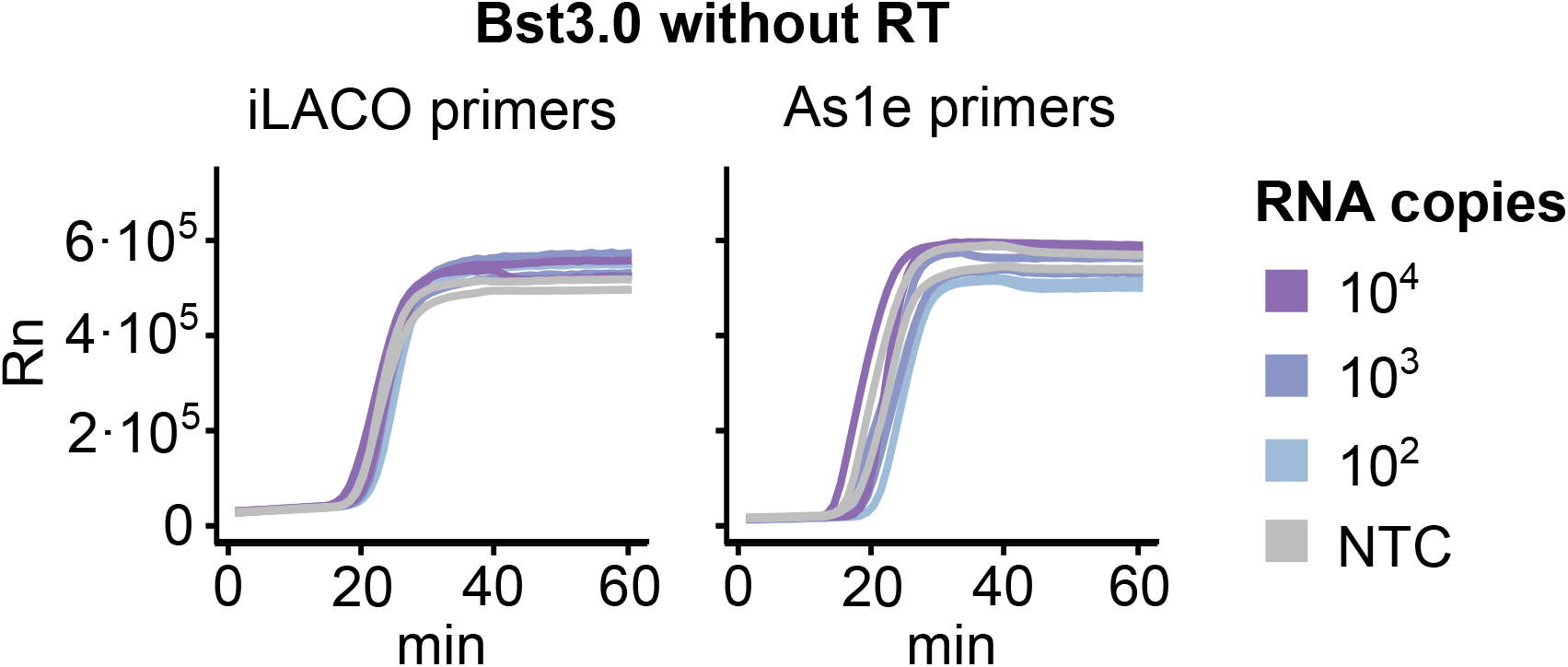
Dependence of Bst3.0 on additional RT activity. Amplification of As1e and iLACO synthetic templates with the corresponding primers using Bst3.0 without an RT enzyme. (Same experiment as Figure 3.)

**Supplementary Table 1**. Contains all the Ct values for clinical samples from GeneXpert and all the RT-LAMP trials.

